# Morphological classes of anemia are associated with hematological parameters but not parasitic infections among women of reproductive age in Kwale, coastal Kenya

**DOI:** 10.1101/2025.09.23.25336509

**Authors:** Lucy Nyambura, Jimmy Hussein Kihara, Francis Mutuku, Moses Ngari, Lydia Kaduka, Collins Okoyo, Charles Mwandawiro, Janet Masaku, Joyce Kamau, Henry Kanyi, Zipporah Ng’ang’a, Victor Tunje Jeza

## Abstract

Anemia disproportionately affects women of reproductive age (WRA) and children below the age of five. Iron deficiency is the major cause of anemia followed by other underlying causes such as malaria, chronic kidney disease, and neglected tropical diseases. This study aimed to determine the morphological classes of anemia and their association with schistosomiasis, malaria, and soil transmitted Helminthes (STH) among WRA in Kwale county, Kenya. Using a cross-sectional study design, parasitic infections and anemia were analyzed from 534 WRA. Frequencies and proportions with 95% confidence intervals were determined. Multilevel mixed-effect logistic regression models were performed to identify variables associated with morphological classes of anemia. The prevalence of normocytic, microcytic and macrocytic anemia was 54%, 43% and 2.3% respectively. The prevalence of Schistosomiasis among women with Normocytic and Microcytic anemia was 4.2% and 3.5%, respectively (P-value=0.72). Microcytic (adjusted Odds Ratio (aOR) 0.65 (95%CI 0.15‒2.78)) compared to normocytic were not associated with Schistosomiasis. The prevalence of malaria among women with Normocytic, Microcytic, and Macrocytic was 5.2%, 4.4%, and 8.3% respectively (P-value=0.78). Microcytic (aOR 1.76 (95%CI 0.43‒7.23)) and macrocytic (aOR 1.75 (95%CI 0.11‒28.0)) compared to normocytic were not associated with malaria. The prevalence of STH among women with Normocytic, Microcytic, and Macrocytic was 7.0%, 3.9%, and 8.3% respectively (P-value=0.31). Microcytic (aOR 0.51 (95%CI 0.13‒2.01)) and macrocytic (aOR 0.72 (95%CI 0.06‒9.39)) compared to normocytic were not associated with STH. Anemia severity (P-value<0.001), RBCs (P-value=0.0001), HCT (P-value=0.0001), platelets (P-value=0.006) MCH (P-value=0.0001) and MCHC (P-value=0.0001) were significantly different across the morphological classes of anemia. In conclusion, the prevalences of normocytic and microcytic anemias were high but not associated with parasitic infections, indicating the possibility of other contributing factors for these morphological classes of anemia in this population.

## INTRODUCTION

A condition known as anemia, or low hemoglobin concentration, occurs when the red blood cells (RBCs) are insufficient and/or their ability to carry enough oxygen to meet the body’s physiological requirements is significantly reduced leading to a state of tissue hypoxia (1,2). It’s a global public health problem affecting some 1.92 billion people worldwide, a sharp increase over the last three decades (3). Reduced iron reserves in the human body leads to low production of hemoglobin thus hindering oxygen from being transported to all organs in the body (4).

There are various methods for classifying anemia (5). Based on hemoglobin concentration, anemia can be classified as none anemic, mild, moderate, or severe while based on RBCs size, anemia can be classified as normocytic, microcytic, or macrocytic. Women of reproductive age and children below the age of five are disproportionately affected by anemia than men (3,6,7). In Kenya, anemia frequency was estimated at 28.7% nationally in 2019 (World Bank, 2021). In addition, a facility based study in Ethiopia found that morphological classes of anemia decreased with increase in age among children (8). Other studies have shown that normocytic anemia was more common followed by microcytic which in turn was more common than macrocytic anemia among women (6).

Schistosomiasis is a neglected tropical disease (NTD) that affects humans after they interact with water contaminated with snails that harbor schistosome cercariae. *Schistosoma haematobium*, *Schistosoma mansoni* and *Schistosoma japonicum* are the three principal trematodes (blood flukes) that cause schistosomiasis in people all over the world. The adult schistosomes, after invading the blood vessel of humans, release thousands of eggs every day through excretion (9). According to WHO global Schistosomiasis atlas of 2016, two hundred million people worldwide were infected by Schistosomiasis and predisposing seven hundred million people. Around the world, *S. japonicum, S. haematobium*, and/or *S. mansoni* infections affect about 40 million women of reproductive age (10). In Kwale, the prevalence of *S. haematobium* among adults was 36.94% among pregnant women and 33.47% among WRA in in 2011 (11), and 3.8% among WRA in 2019 (12), a significant drop that was attributed majorly to mass drug administration through the TUMIKIA project, school based deworming (SBD) campaigns, and use of improved water, sanitation, and hygiene (WASH) factors (12,13).

The most common Soil Transmitted Helminthes affecting majorly low economic status communities in the sub-Saharan Africa region include round worms (*Ascaris Lumbricoides*) whipworms (*Trichuris Trichiura*), and hookworms (*Anyclostoma duodenal*). A study done in Nigeria found that intestinal helminthes infections were significantly associated with anemia burden among children (14). Although deworming in schools –school based deworming (SBD) program was considered to have lowered the incidence of STH infections, this did not reduce or stop transmission at the community level. Many studies have proven that combining community based deworming (CBD) and SBD may be effective in breaking transmission (13). However, excessive use of anti-helminthes drugs have been shown to drive single nucleotide polymorphism pressure leading to drug resistance (15). To counter this, integrated approaches to combating helminthes infections have been proposed to protect current and future anti-helminthes drug resistance (16).

Malaria disease is more frequent among the poor and contributes to poverty. In Kenya, it is more prevalent in the western and coastal regions. In Kwale, the prevalence of malaria was 4.9% among WRA in 2019 (12), and it was estimated to be 6% nationwide among children from six months to fourteen years old in 2020 (17).

Iron deficiency is the major cause of anemia followed by other underlying causes such as malaria, chronic kidney disease, and neglected tropical diseases (NTDs) (3). These NTDs include the aforementioned schistosomiasis and STH infections. Although the exact mechanisms linking schistosomiasis and anemia remain unclear, plausible explanations include hemolysis, inflammatory processes, and bone marrow suppression (18). A previous hypothesis was that schistosomiasis could contribute to iron deficiency anemia due to extracorporeal blood loss; splenic sequestration, autoimmune hemolysis and anemia of inflammation (AoI) (19). In Western Kenya, AoI was most commonly associated with *S. mansoni* infections, contrasting with iron deficiency anemia in uninfected children. The prevalence of AoI showed a significant trend with the intensity of *S. mansoni* infection (20). Additionally, soil-transmitted helminth infection contributes to anemia by causing iron shortage and vitamin depletion. Many studies have been done looking at schistosomiasis, STH infections, and malaria and their association with anemia. For example, a meta-analysis by Kassebaum *et. al.* identified malaria and schistosomiasis as leading contributors to the prevalence of anemia (21). Further, a research by Salgado and others found a correlation between *Schistosoma mansoni* infection with serum ferritin levels but not hemoglobin, even after adjusting for malaria parasitemia and hookworm (22). In addition, a recent research of pregnant women receiving antenatal care (ANC) at Msambweni Hospital evaluated the prevalence and risk factors for anemia in pregnancy and malaria in pregnancy (23). The study discovered a 62.7% prevalence of anemia in pregnancy, which was linked to, among other things, malaria parasitemia, being in the third trimester of pregnancy, and having a passion for eating soil while pregnant (23). Although the study highlights the prevalence of anemia and some of its correlates, the study was conducted in one hospital that is a county referral hospital and did not explore the effect/associations of morphological classes of anemia. To the best of our knowledge, no studies have been done to determine the association of parasitic infections with morphological classes of anemia in Kwale. The current study sought to fill this gap.

## MATERIALS AND METHODS

### Study Design, site, sample size, and Socio-demographic characteristics

The design, site, population, and socio-demographic characteristics of the study are explained in detail elsewhere (12). Briefly, this was a cross-sectional study design conducted in Kwale, south coast of Kenya involving 534 WRA between 15 and 50 years old. Sample and data collection for this reported study was done from last week of November to the first week of December, 2018. Socio-demographic characteristics were collected using a questionnaire originally developed in English and later translated to Swahili language which was more understandable for most of the participants. The translation was then done backwards to English to ensure the same meaning was maintained.

### Parasitological examinations

#### S. haematobium infection

This was determined by examining fresh urine samples using nuclear pore filtration technique. To do this, 10 ml of urine was collected from WRA between 10 am and 2 pm to coincide with the peak production of eggs by the blood fluke *S. haematobium*. The urine was then filtered in duplicate through 12.0 µm (13 mm) pore-sized polycarbonate membrane filters (Sterlitech, Kent, WA, USA) mounted on urine filtration chambers. The membranes were then placed on labeled slides and examined using a microscope under power 40X. Mean egg counts were calculated and expressed as egg counts per 10 ml of urine.

#### Soil Transmitted Helminths infection

The presence or absence of STH (*Trichiuriasis, Ascariasis* and hookworm infections) ova in stool was analyzed by Kato-Katz method (24). Briefly, duplicate thick smears were prepared using a sieve and a calibrated template designed to hold 41.7 mg stool. The duplicate smears from each participant were placed onto glass slides and covered with cellophane strips pre-soaked in glycerol-malachite green solution. The slides were examined for STH ova using a microscope within one hour of preparation. Egg counts were multiplied by a factor of 24 to obtain the eggs per gram of stool.

#### Blood slides for malaria parasites

A drop of venous blood was used to make thick and thin smears. These were then stained with 2% Giemsa solution for 30 minutes. Then, gently the stain was flushed off the slide by adding drops of clean-buffered distilled or deionized water at pH 7.0 for one minute. The slides were placed film side downward in a slide rack to drain and dry, making sure the film does not touch the rack. The slides were examined using a compound microscope - 100x power and malaria parasites identified (deep red chromatin and pale purplish blue cytoplasm). A blood smear with a negative result, denoted none existence of asexual parasites. Gametocytes were also determined from the thick blood smears. Thin smears were used to differentiate the different Plasmodium species. For quality control, 10% of all the slides were read again.

### Analysis for Anemia by Full Blood Count test

Anemia was analyzed using a full blood count test done by a haematology analyzer (Nihon Kohden Celltac MEK8222). Anemia was analyzed based on mean corpuscular volume (MCVs) obtained from the full blood count and categorized as Normocytic, Microcytic, and Macrocytic. MCV has survived as a key parameter for the classification of anemia with automated hematology analyzers for almost 100 years since Maxwel Wintrobe proposed classification of anemia based on the mean cell volume (25). Based on hemoglobin concentration, anemia was further classified by severity as none, mild, moderate, and severe.

### Data Processing and Analysis

The prevalence of Schistosomiasis, Malaria, Soil Transmitted Helminthes and the 3 morphological classes of anemia were reported as proportions with binomial exact 95% confidence intervals. The prevalence of Schistosomiasis, Malaria, and STH between/across selected variables like age group, sub-county, pregnancy and WASH characteristics were compared using chi-square/fisher’s exact tests. The prevalence of the morphological classes of anemia across/between categorical variables were compared using chi-square/fisher’s exact tests while Kruskal–Wallis test, a non-parametric test, was used to compare medians of continuous laboratory parameters across the three levels of morphological classes of anemia. The laboratory parameters were skewed and therefore could not be compared using parametric tests. To determine the association between the morphological classes of anemia, with schistosomiasis, malaria, and STH, multilevel mixed-effect logistic regression model was fixed with the village as random intercept to account for clustering within the villages. The binary schistosomiasis, malaria, and STH variables (either negative or positive) were the dependent variables in the multilevel mixed-effect logistic regression models. Three regression models were fixed for each dependent variable. In the univariate multilevel mixed-effed logistic regression models, the only independent variable in the model was the morphological classes of anemia variable with normocytic level as the reference. In the multivariable multilevel mixed-effect logistic regression models, we adjusted for identified variables associated with morphological classes of anemia in the univariable analysis (those with a P-value <0.05). However, because hemoglobin levels are on the casual pathways it was not included in the multivariable models. Statistical analyses were conducted using R (version 4.2.0) and STATA version 17.0 (StataCorp, College Station, TX, USA).

### Ethical Considerations

The Scientific and Ethics Review Unit of Kenya Medical Research Institute (Ref No. KEMRI/SERU/ESACIPAC/3684) reviewed and approved the study. Licensing of the study was acquired from the Kenya National Commission for Science, Technology, and Innovation (NACOSTI) in accordance with the rules and regulations that govern all studies in Kenya. Permission to carry out the study in Kwale County was sought through the County Department of Health services. All participants provided a signed consent in Swahili or English after the study was explained to them. For those below 18 years, assent and consent were signed by them and their legal guardians or parents respectively. For WRA who may not have been able to read for any reason, the consent form was read to them and signed by imprinting using the thumb finger. All participants were made aware that they could withdraw from the study at any time. A unique identifier was used to help in tracking the sample of each participant without disclosing the identity of the participants. Further, the questionnaire was masked in order to protect the confidentiality of the participants according to global ethical standards and procedures and filed safely to prevent data leakage or its misuse.

## Results

### The prevalence of morphological classes of anemia among women of reproductive age

There were 285, 228, and 12 women with normocytic, microcytic, and macrocytic morphological classes of anemia which translates to a prevalence of 54.3% (95%CI 49.9‒58.6), 43.4% (95%CI 39.1‒47.8) and 2.3% (95%CI 1.20‒4.0) respectively as shown in **Fig 1**.

**Fig 1.**
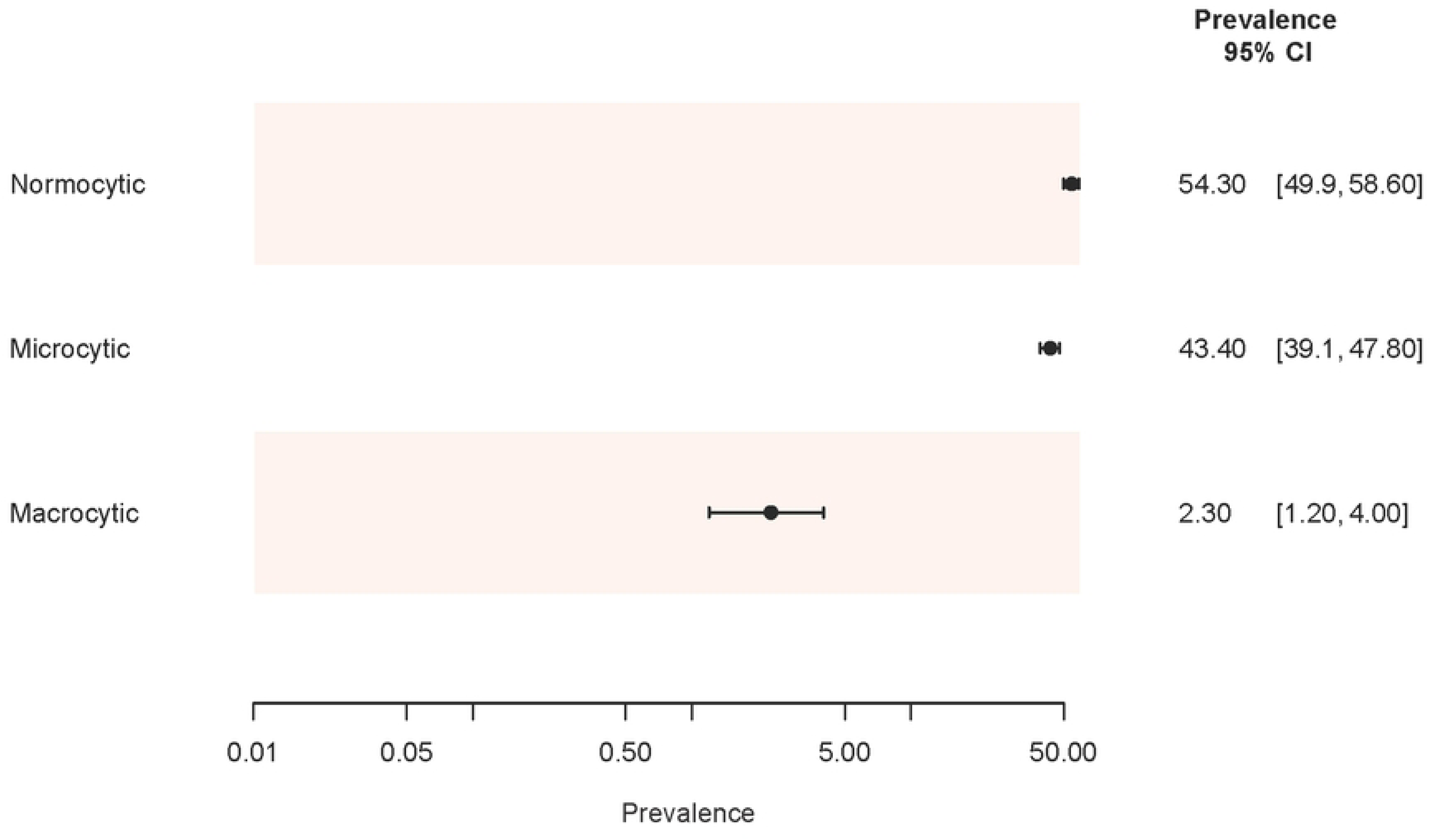
Prevalence of morphological classes of anemia among women of reproductive age.

The prevalence (95% CI) of Schistosomiasis among women with Normocytic and Microcytic anemia was 4.2% (95% CI 2.2‒7.2) and 3.5% (95% CI 1.5‒6.8) respectively (P-value=0.72). No woman with Macrocytic anemia was infected with Schistosomiasis. The prevalence (95% CI) of malaria among women with Normocytic, Microcytic, and Macrocytic was 5.2% (95% CI 3.0‒8.5), 4.4% (95%CI 2.1‒7.9), and 8.3% (95%CI 0.2‒38.5) respectively (P-value=0.78). The prevalence (95% CI) of STH among women with Normocytic, Microcytic, and Macrocytic was 7.0% (95% CI 4.3‒10.6), 3.9% (95% CI 1.8‒7.4), and 8.3% (95% CI 0.2‒38.5) respectively (P-value=0.31) as shown in **Fig 2**.

**Fig 2:**
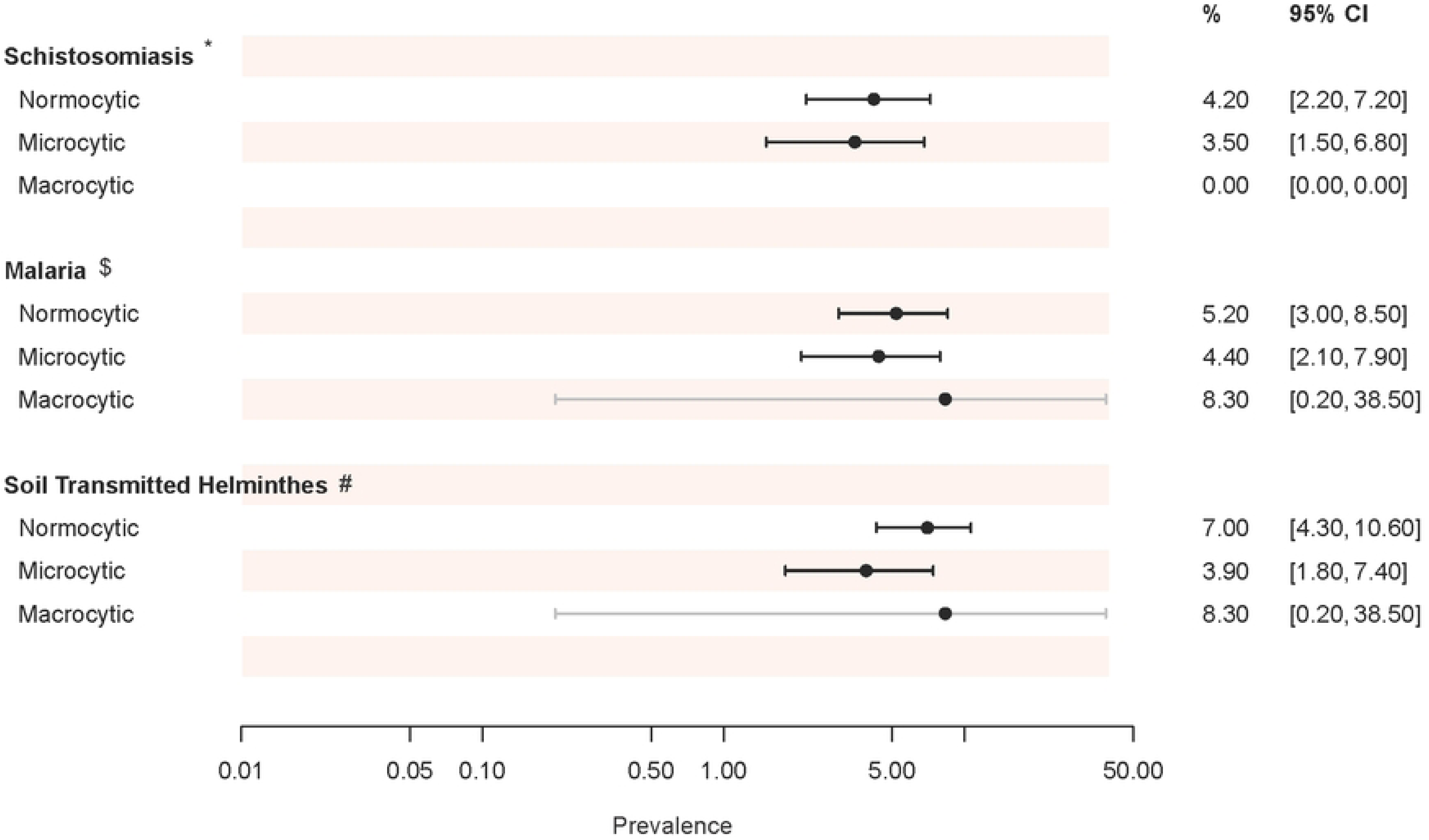
Prevalence of Schistosomiasis, Malaria, and STH across the morphological classes of anemia among women of reproductive age. P-values: *=0.72, $=0.78, #=0.31.

### Association between parasitic infections (Schistosomiasis, Malaria, and STH) and morphological classes of anemia

Age (P-value=0.02), ever pregnant (P-value=0.005), hand wash after toilet use (P-value=0.04), anemia (P-value <0.001), red blood cells (P-value=0.0001), HCT (%) (P-value=0.0001), platelets (P-value=0.006) MCH (P-value=0.0001) and MCHC (P-value=0.0001) were significantly different across the morphological classes of anemia as shown in **Table 1** and **Figure 3**.

**Fig 3.**
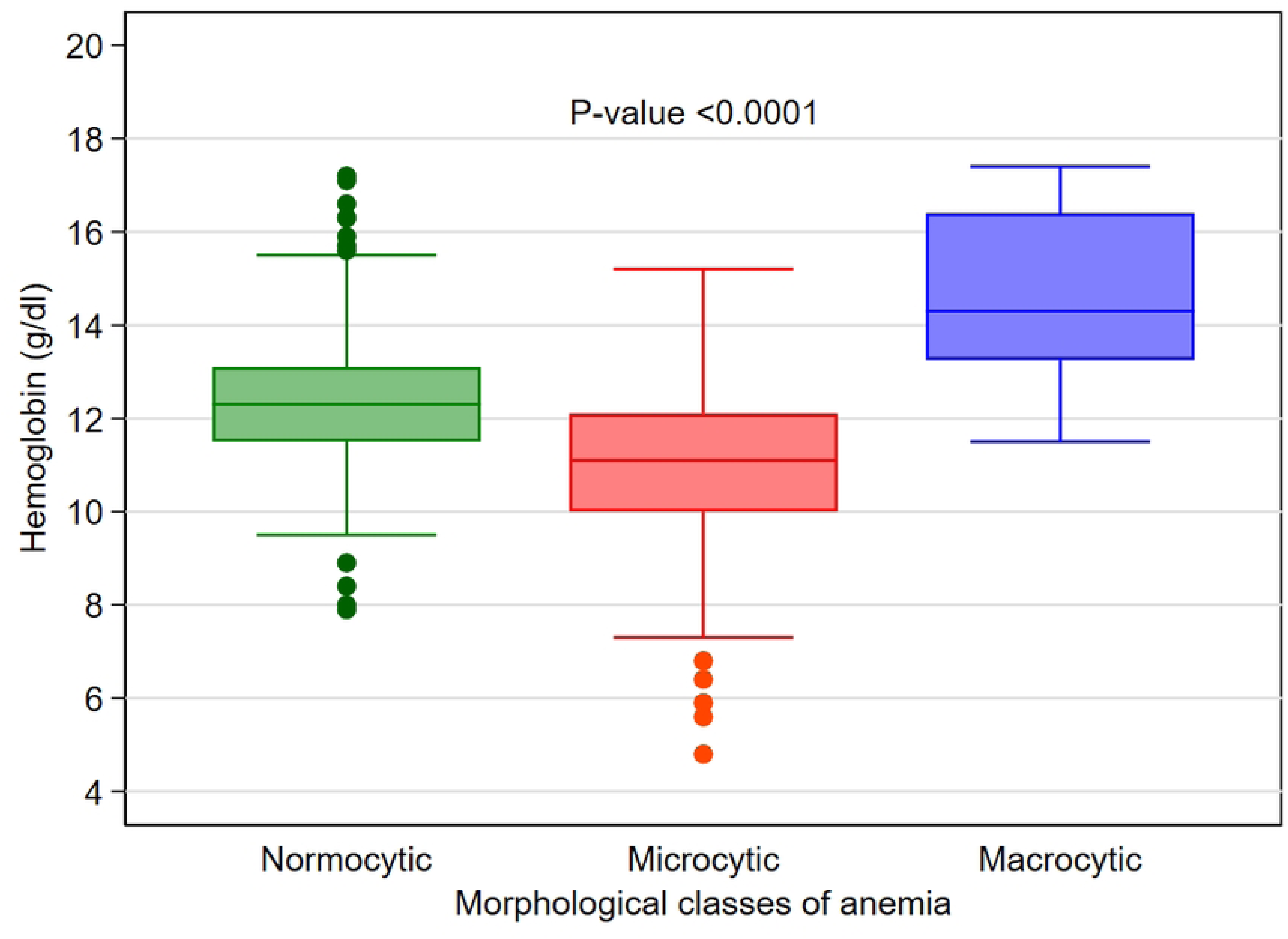
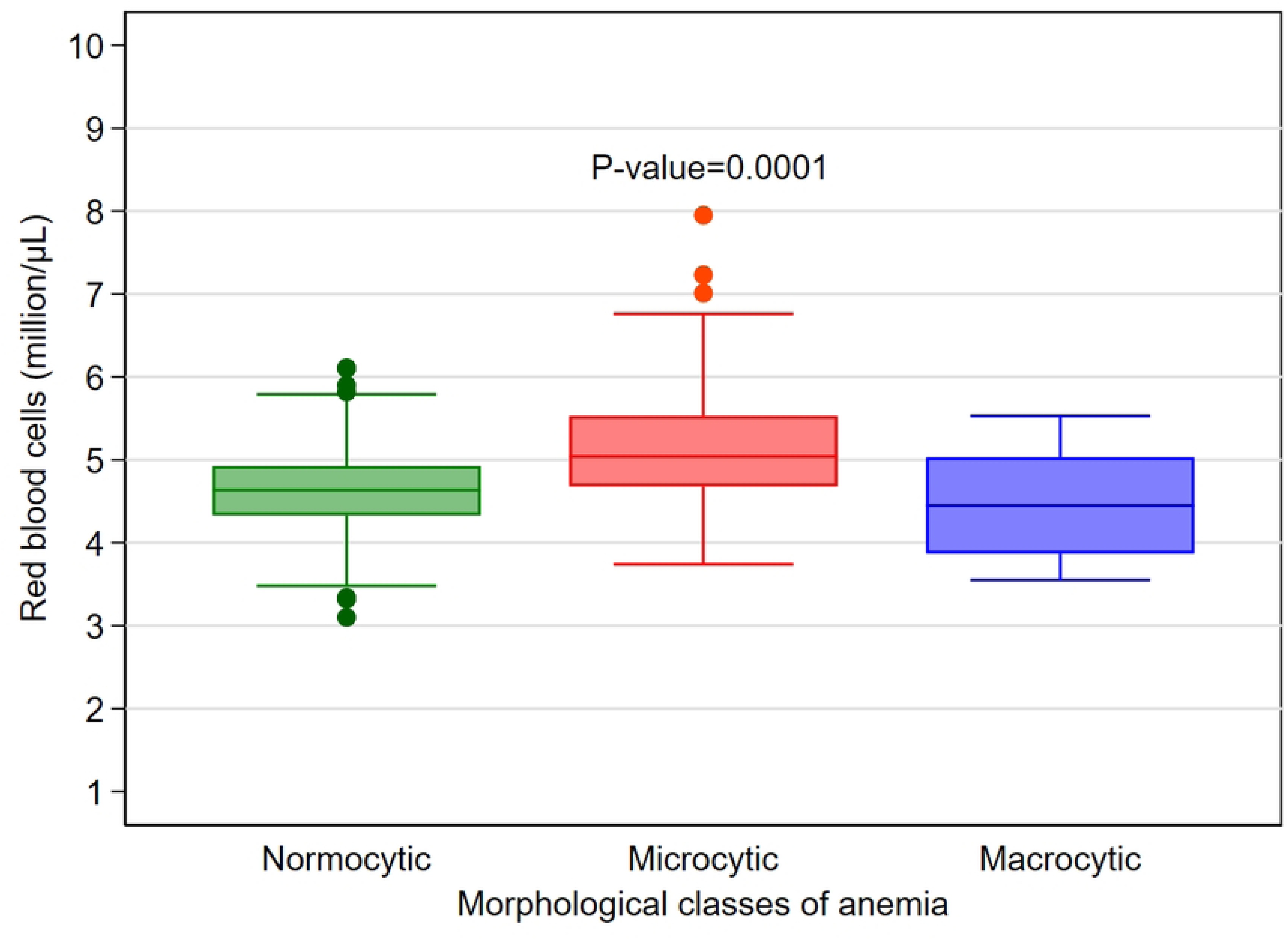
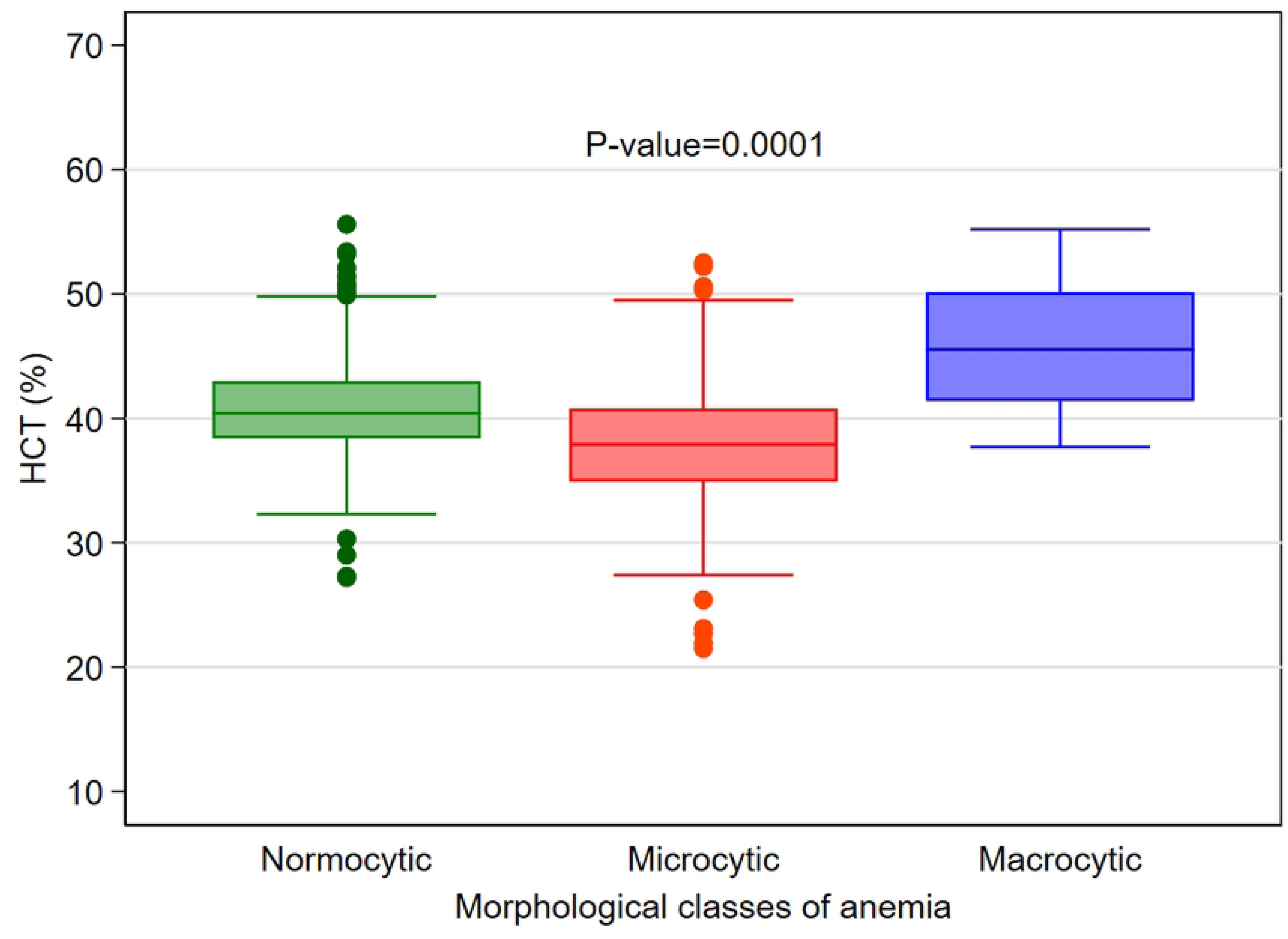
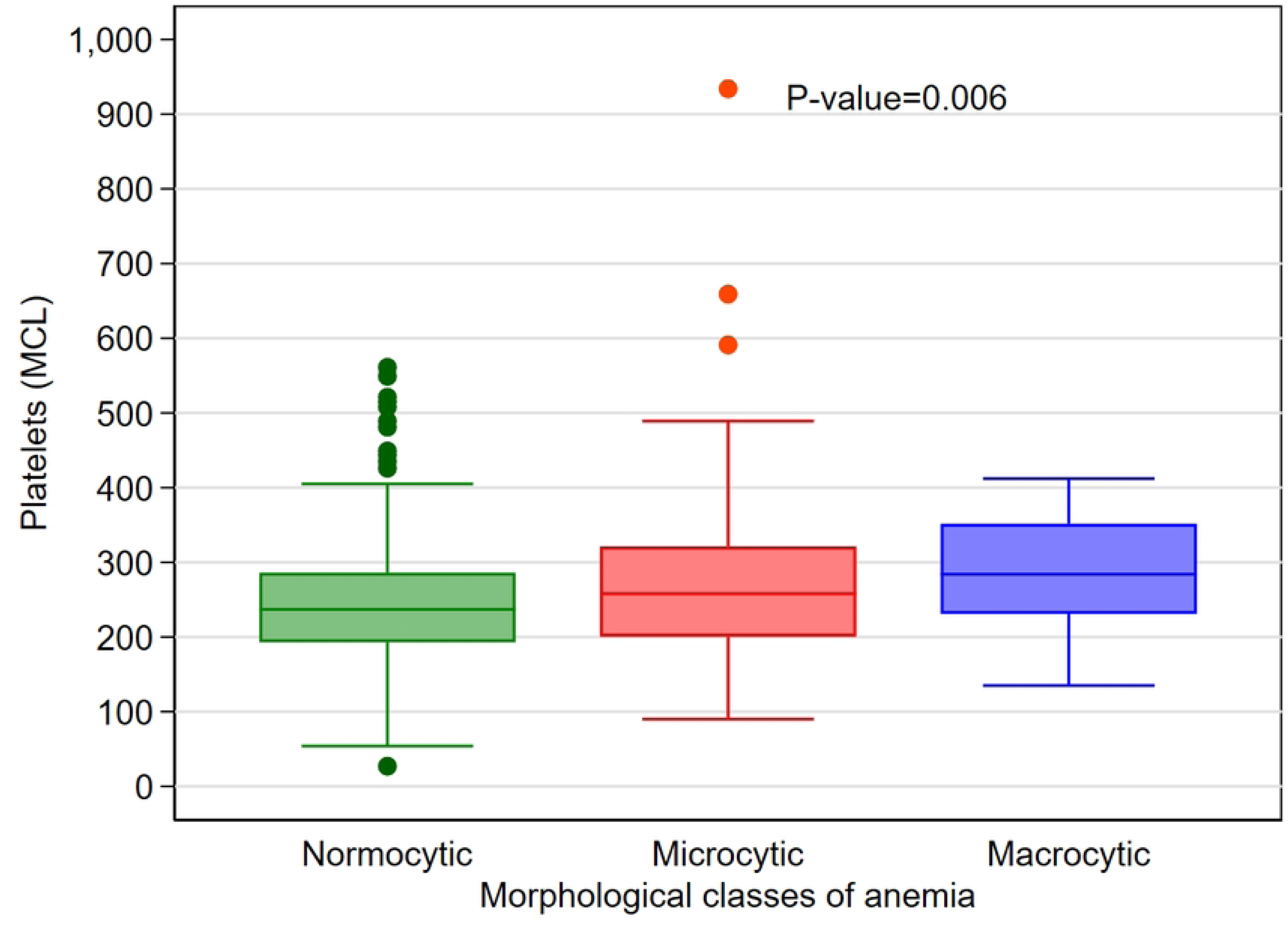
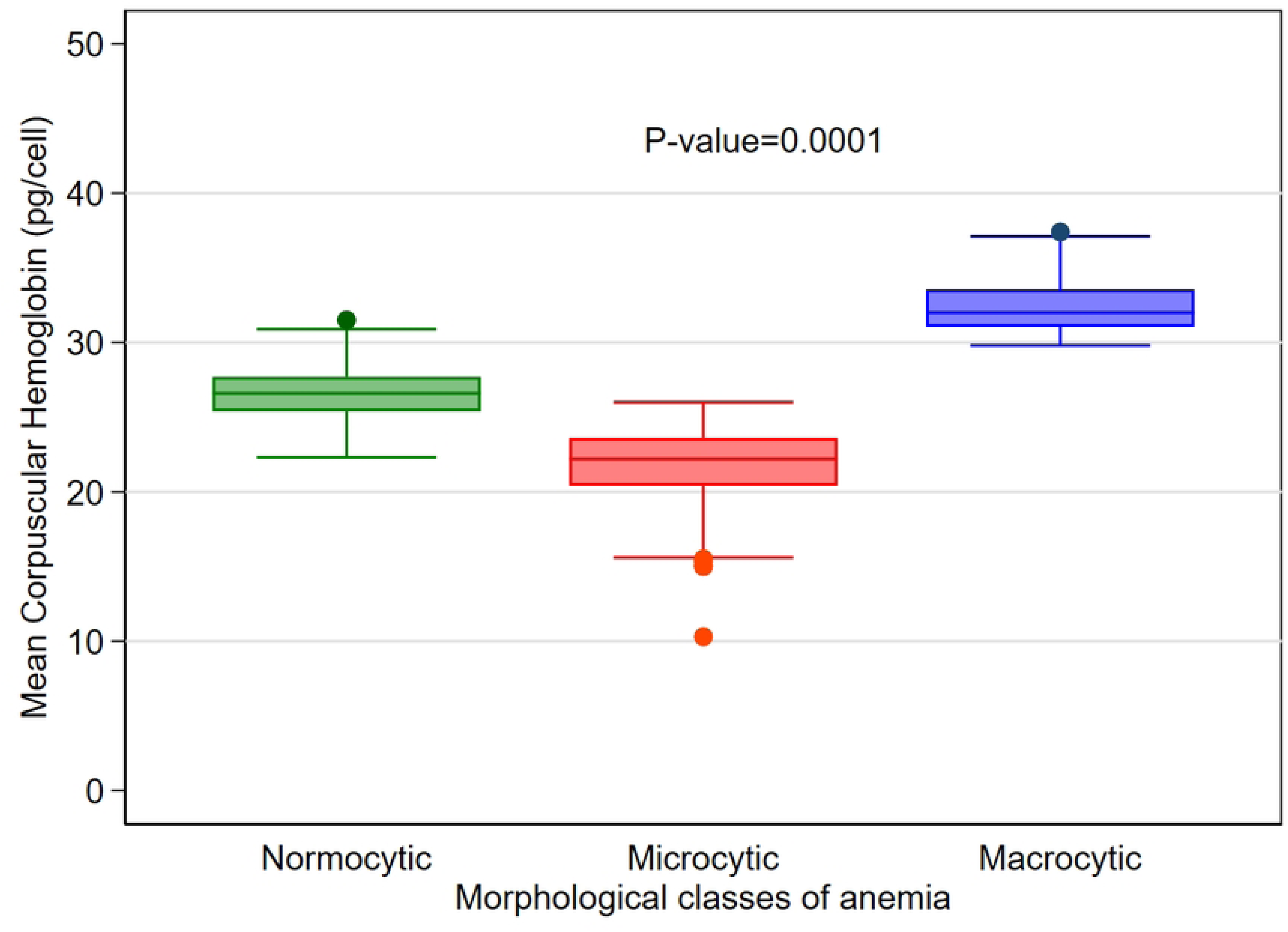
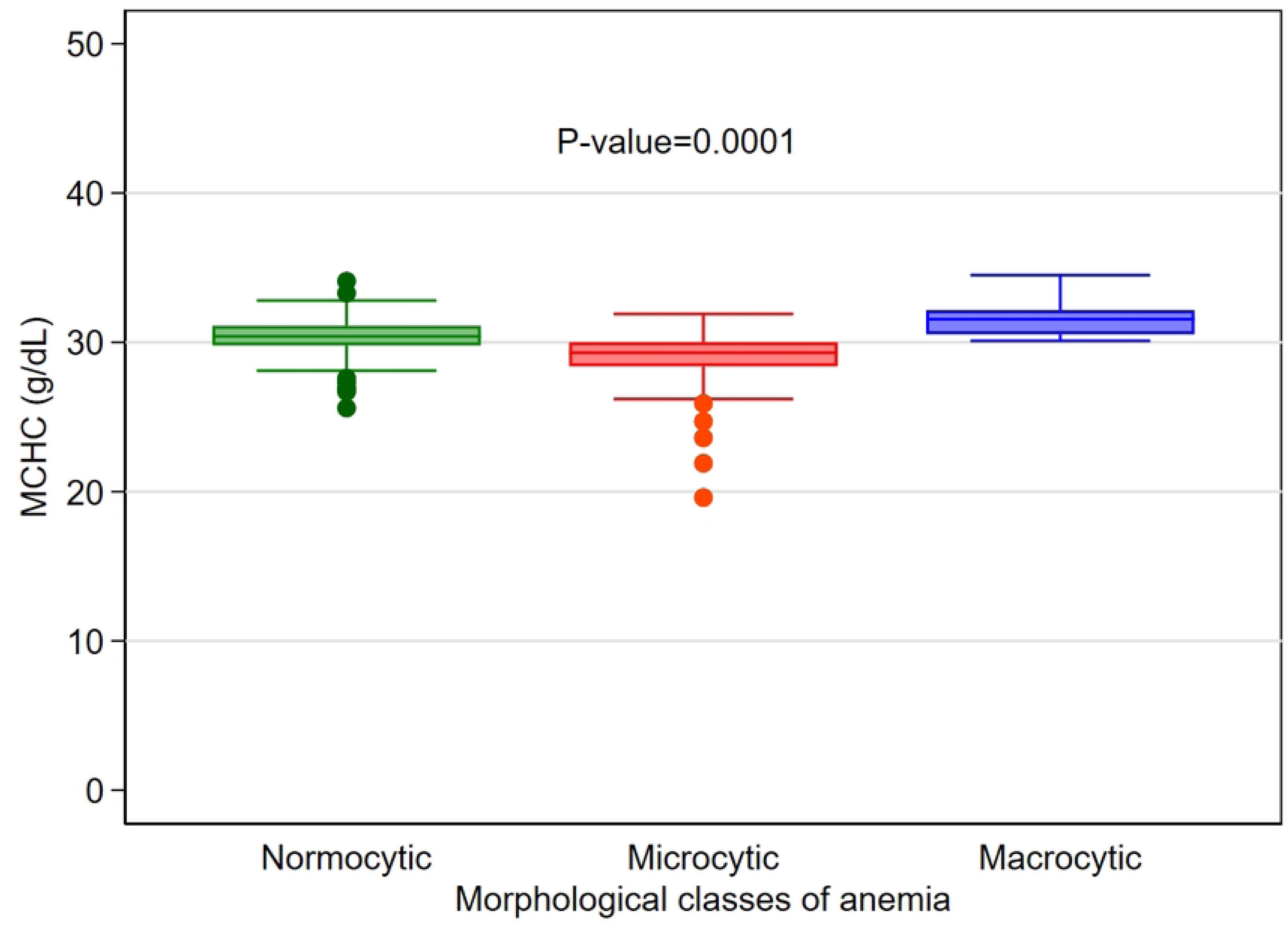
Box plots showing the distribution of: a) Hemoglobin (g/dl); b) Red blood cells; c) HCT (%); d) Platelets; e) MCH and f) MCHC across the morphological classes of anemia. P-values are from Kruskal–Wallis test reported in Table 1 above.

**Table 1.**
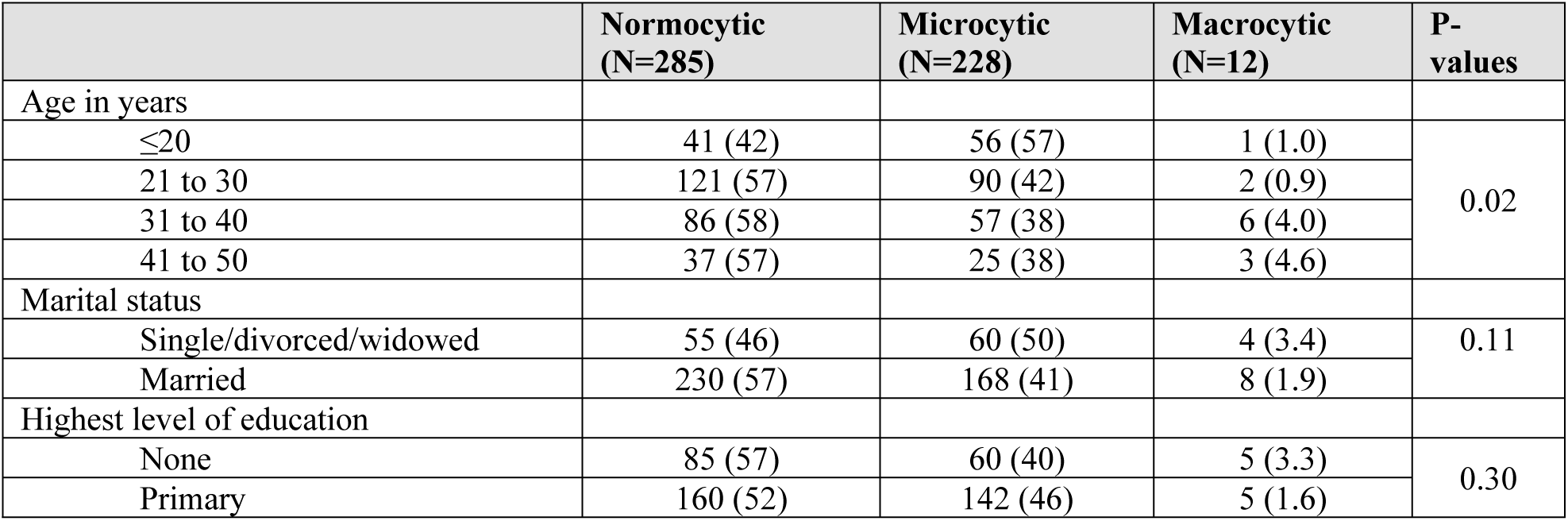
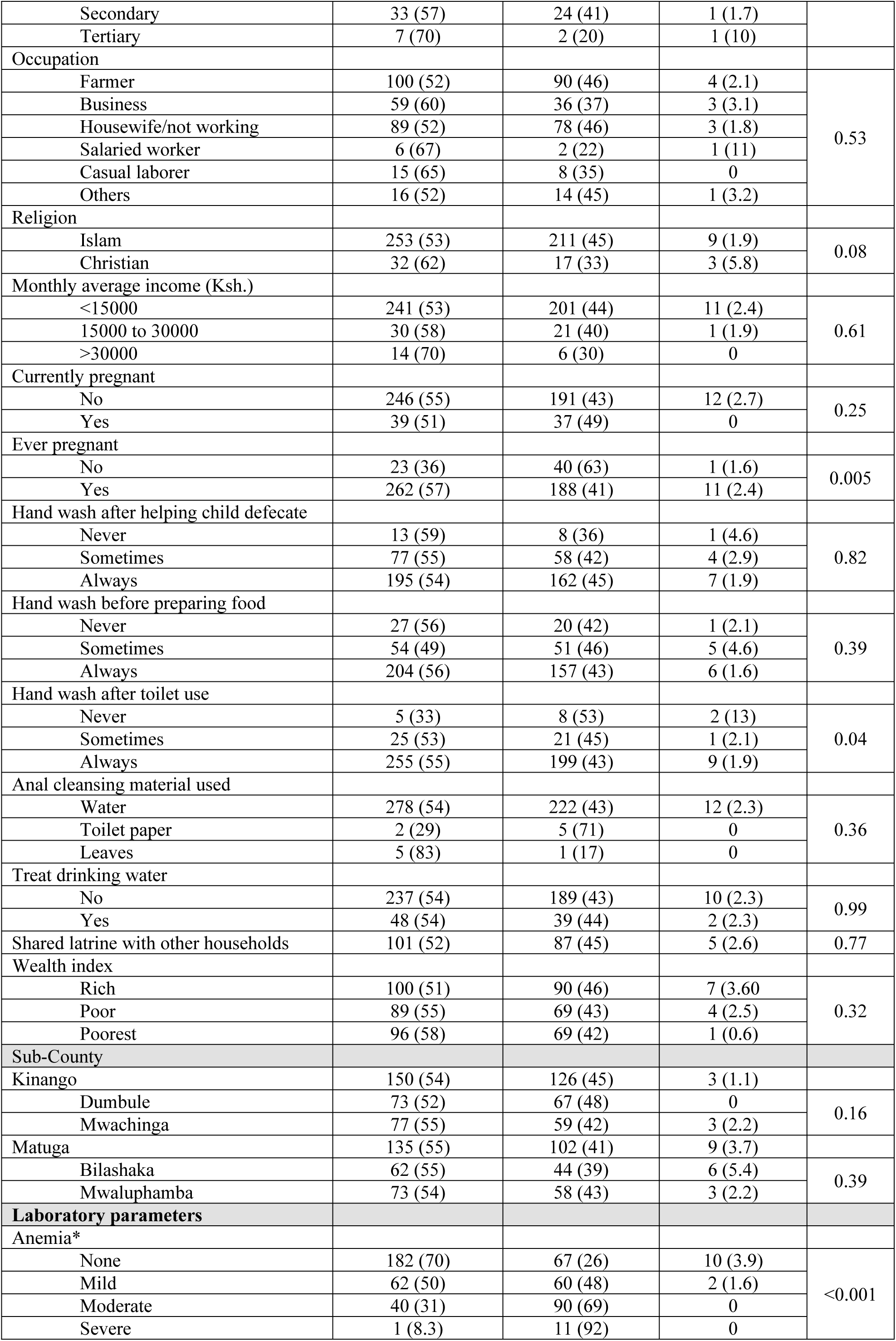
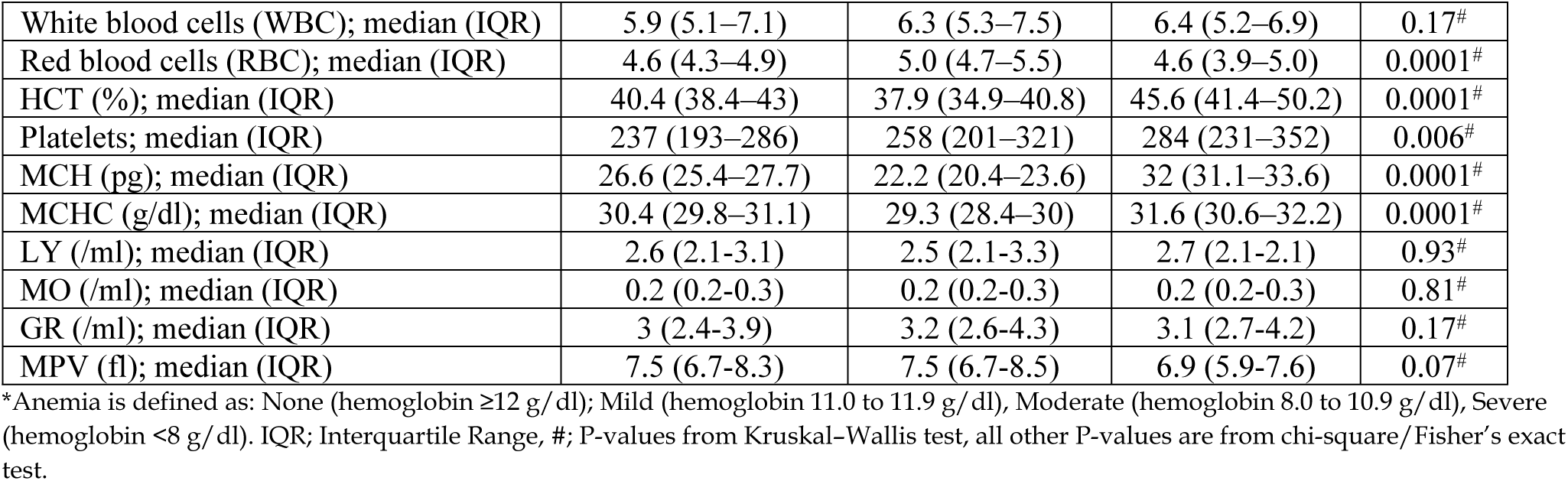
Distribution of select variables across the morphological classes of anemia among women of reproductive age.

|  | Normocytic<br>(N=285) | Microcytic<br>(N=228) | Macrocytic<br>(N=12) | P-values |
| --- | --- | --- | --- | --- |
| Age in years |  |  |  |  |
| ≤20 | 41 (42) | 56 (57) | 1 (1.0) | 0.02 |
| 21 to 30 | 121 (57) | 90 (42) | 2 (0.9) |  |
| 31 to 40 | 86 (58) | 57 (38) | 6 (4.0) |  |
| 41 to 50 | 37 (57) | 25 (38) | 3 (4.6) |  |
| Marital status |  |  |  |  |
| Single/divorced/widowed | 55 (46) | 60 (50) | 4 (3.4) | 0.11 |
| Married | 230 (57) | 168 (41) | 8 (1.9) |  |
| Highest level of education |  |  |  |  |
| None | 85 (57) | 60 (40) | 5 (3.3) | 0.30 |
| Primary | 160 (52) | 142 (46) | 5 (1.6) |  |
| Secondary | 33 (57) | 24 (41) | 1 (1.7) |  |
| Tertiary | 7 (70) | 2 (20) | 1 (10) |  |
| Occupation |  |  |  |  |
| Farmer | 100 (52) | 90 (46) | 4 (2.1) | 0.53 |
| Business | 59 (60) | 36 (37) | 3 (3.1) |  |
| Housewife/not working | 89 (52) | 78 (46) | 3 (1.8) |  |
| Salaried worker | 6 (67) | 2 (22) | 1 (11) |  |
| Casual laborer | 15 (65) | 8 (35) | 0 |  |
| Others | 16 (52) | 14 (45) | 1 (3.2) |  |
| Religion |  |  |  |  |
| Islam | 253 (53) | 211 (45) | 9 (1.9) | 0.08 |
| Christian | 32 (62) | 17 (33) | 3 (5.8) |  |
| Monthly average income (Ksh.) |  |  |  |  |
| <15000 | 241 (53) | 201 (44) | 11 (2.4) | 0.61 |
| 15000 to 30000 | 30 (58) | 21 (40) | 1 (1.9) |  |
| >30000 | 14 (70) | 6 (30) | 0 |  |
| Currently pregnant |  |  |  |  |
| No | 246 (55) | 191 (43) | 12 (2.7) | 0.25 |
| Yes | 39 (51) | 37 (49) | 0 |  |
| Ever pregnant |  |  |  |  |
| No | 23 (36) | 40 (63) | 1 (1.6) | 0.005 |
| Yes | 262 (57) | 188 (41) | 11 (2.4) |  |
| Hand wash after helping child defecate |  |  |  |  |
| Never | 13 (59) | 8 (36) | 1 (4.6) | 0.82 |
| Sometimes | 77 (55) | 58 (42) | 4 (2.9) |  |
| Always | 195 (54) | 162 (45) | 7 (1.9) |  |
| Hand wash before preparing food |  |  |  |  |
| Never | 27 (56) | 20 (42) | 1 (2.1) | 0.39 |
| Sometimes | 54 (49) | 51 (46) | 5 (4.6) |  |
| Always | 204 (56) | 157 (43) | 6 (1.6) |  |
| Hand wash after toilet use |  |  |  |  |
| Never | 5 (33) | 8 (53) | 2 (13) | 0.04 |
| Sometimes | 25 (53) | 21 (45) | 1 (2.1) |  |
| Always | 255 (55) | 199 (43) | 9 (1.9) |  |
| Anal cleansing material used |  |  |  |  |
| Water | 278 (54) | 222 (43) | 12 (2.3) | 0.36 |
| Toilet paper | 2 (29) | 5 (71) | 0 |  |
| Leaves | 5 (83) | 1 (17) | 0 |  |
| Treat drinking water |  |  |  |  |
| No | 237 (54) | 189 (43) | 10 (2.3) | 0.99 |
| Yes | 48 (54) | 39 (44) | 2 (2.3) |  |
| Shared latrine with other households | 101 (52) | 87 (45) | 5 (2.6) | 0.77 |
| Wealth index |  |  |  |  |
| Rich | 100 (51) | 90 (46) | 7 (3.60) | 0.32 |
| Poor | 89 (55) | 69 (43) | 4 (2.5) |  |
| Poorest | 96 (58) | 69 (42) | 1 (0.6) |  |
| Sub-County |  |  |  |  |
| Kinango | 150 (54) | 126 (45) | 3 (1.1) | 0.16 |
| Dumbule | 73 (52) | 67 (48) | 0 |  |
| Mwachinga | 77 (55) | 59 (42) | 3 (2.2) |  |
| Matuga | 135 (55) | 102 (41) | 9 (3.7) | 0.39 |
| Bilashaka | 62 (55) | 44 (39) | 6 (5.4) |  |
| Mwaluphamba | 73 (54) | 58 (43) | 3 (2.2) |  |
| **Laboratory parameters** |  |  |  |  |
| Anemia* |  |  |  |  |
| None | 182 (70) | 67 (26) | 10 (3.9) | <0.001 |
| Mild | 62 (50) | 60 (48) | 2 (1.6) |  |
| Moderate | 40 (31) | 90 (69) | 0 |  |
| Severe | 1 (8.3) | 11 (92) | 0 |  |
| White blood cells (WBC); median (IQR) | 5.9 (5.1–7.1) | 6.3 (5.3–7.5) | 6.4 (5.2–6.9) | 0.17<sup>#</sup> |
| Red blood cells (RBC); median (IQR) | 4.6 (4.3–4.9) | 5.0 (4.7–5.5) | 4.6 (3.9–5.0) | 0.0001<sup>#</sup> |
| HCT (%); median (IQR) | 40.4 (38.4–43) | 37.9 (34.9–40.8) | 45.6 (41.4–50.2) | 0.0001<sup>#</sup> |
| Platelets; median (IQR) | 237 (193–286) | 258 (201–321) | 284 (231–352) | 0.006<sup>#</sup> |
| MCH (pg); median (IQR) | 26.6 (25.4–27.7) | 22.2 (20.4–23.6) | 32 (31.1–33.6) | 0.0001<sup>#</sup> |
| MCHC (g/dl); median (IQR) | 30.4 (29.8–31.1) | 29.3 (28.4–30) | 31.6 (30.6–32.2) | 0.0001<sup>#</sup> |
| LY (/ml); median (IQR) | 2.6 (2.1–3.1) | 2.5 (2.1–3.3) | 2.7 (2.1–2.1) | 0.93<sup>#</sup> |
| MO (/ml); median (IQR) | 0.2 (0.2–0.3) | 0.2 (0.2–0.3) | 0.2 (0.2–0.3) | 0.81<sup>#</sup> |
| GR (/ml); median (IQR) | 3 (2.4–3.9) | 3.2 (2.6–4.3) | 3.1 (2.7–4.2) | 0.17<sup>#</sup> |
| MPV (fl); median (IQR) | 7.5 (6.7–8.3) | 7.5 (6.7–8.5) | 6.9 (5.9–7.6) | 0.07<sup>#</sup> |

\*Anemia is defined as: None (hemoglobin $\geq 12$ g/dl); Mild (hemoglobin 11.0 to 11.9 g/dl), Moderate (hemoglobin 8.0 to 10.9 g/dl), Severe (hemoglobin $< 8$ g/dl). IQR; Interquartile Range, #; P-values from Kruskal-Wallis test, all other P-values are from chi-square/Fisher's exact test.

### The association between the morphological classes of anemia, with schistosomiasis, malaria, and Soil Transmitted Helminthes

In the univariate analysis, Microcytic Crude Odds Ratio (COR) 0.83 (95%CI 0.33‒2.06)) compared to normocytic were not associated with Schistosomiasis. After adjusting for potential confounders, Microcytic Adjusted Odds Ratio (aOR) 0.65 (95%CI 0.15‒2.78)) compared to normocytic were not associated with Schistosomiasis as shown in **Table 2**.

**Table 2.** Analysis of the association between the morphological classes of anemia, with schistosomiasis, among women of reproductive age.

|  | Schistosomiasis |  | Multivariable analysis |  |
| --- | --- | --- | --- | --- |
|  | No (N=505) | Yes (N=20) | Adjusted Odds Ratio (95%CI) | P-value |
|  | Normocytic | Microcytic | Macrocytic |  |
| Morphological classes of anemia |  |  |  |  |
| Normocytic | 273 (96) | 12 (4.2) | Reference |  |
| Microcytic | 220 (96) | 8 (3.5) | 0.65 (0.15–2.78) | 0.56 |
| Macrocytic | 12 (100) | 0 | - |  |
| Age in years |  |  |  |  |
| $\leq 20$ | 92 (94) | 6 (6.1) | Reference | |
| 21 to 30 | 202 (95) | 11 (5.2) | 0.72 (0.21–2.42) | 0.59 |
| 31 to 40 | 146 (98) | 3 (2.0) | 0.27 (0.06–1.30) | 0.10 |
| 41 to 50 | 65 (100) | 0 | - |  |
| Ever pregnant |  |  |  |  |
| No | 61 (95) | 3 (4.7) | Reference |  |
| Yes | 444 (96) | 17 (3.7) | 0.79 (0.17–3.63) | 0.77 |
| Hand wash after toilet use |  |  |  |  |
| Never | 15 (100) | 0 | - |  |
| Sometimes | 45 (96) | 2 (4.3) | 1.08 (0.23–4.97) | 0.92 |
| Always | 445 (96) | 18 (3.9) | Reference |  |
| Red blood cells (RBC) | 4.8 (4.4–5.2) | 4.7 (4.3–5.2) | 0.91 (0.11–7.63) | 0.93 |
| HCT (%) | 39.6 (36.9–42.4) | 38.1 (35.3–41.3) | 0.99 (0.75–1.31) | 0.95 |
| Platelets | 247 (197–302) | 237 (183–327) | 0.99 (0.97–1.03) | 0.89 |
| MCH (pg) | 25 (22.4–27.1) | 24.9 (22.7–26.3) | 1.01 (0.63–1.63) | 0.97 |
| MCHC (g/dl) | 30 (29.2–30.7) | 29.7 (29.1–30.2) | 0.79 (0.50–1.25) | 0.30 |

Adjusted Odds Ratio and P-values are from multilevel logistic regression analysis.

In the univariate analysis, Microcytic (COR 0.83 (95%CI 0.36‒1.90)) and Macrocytic (COR 1.53 (95%CI 0.18‒13.2)) compared to normocytic were not associated with malaria. After adjusting for potential confounders, Microcytic (aOR 1.76 (95%CI 0.43‒7.23)) and macrocytic (aOR 1.75 (95%CI 0.11‒28.0)) compared to normocytic were not associated with malaria as shown in **Table 3**.

**Table 3.**
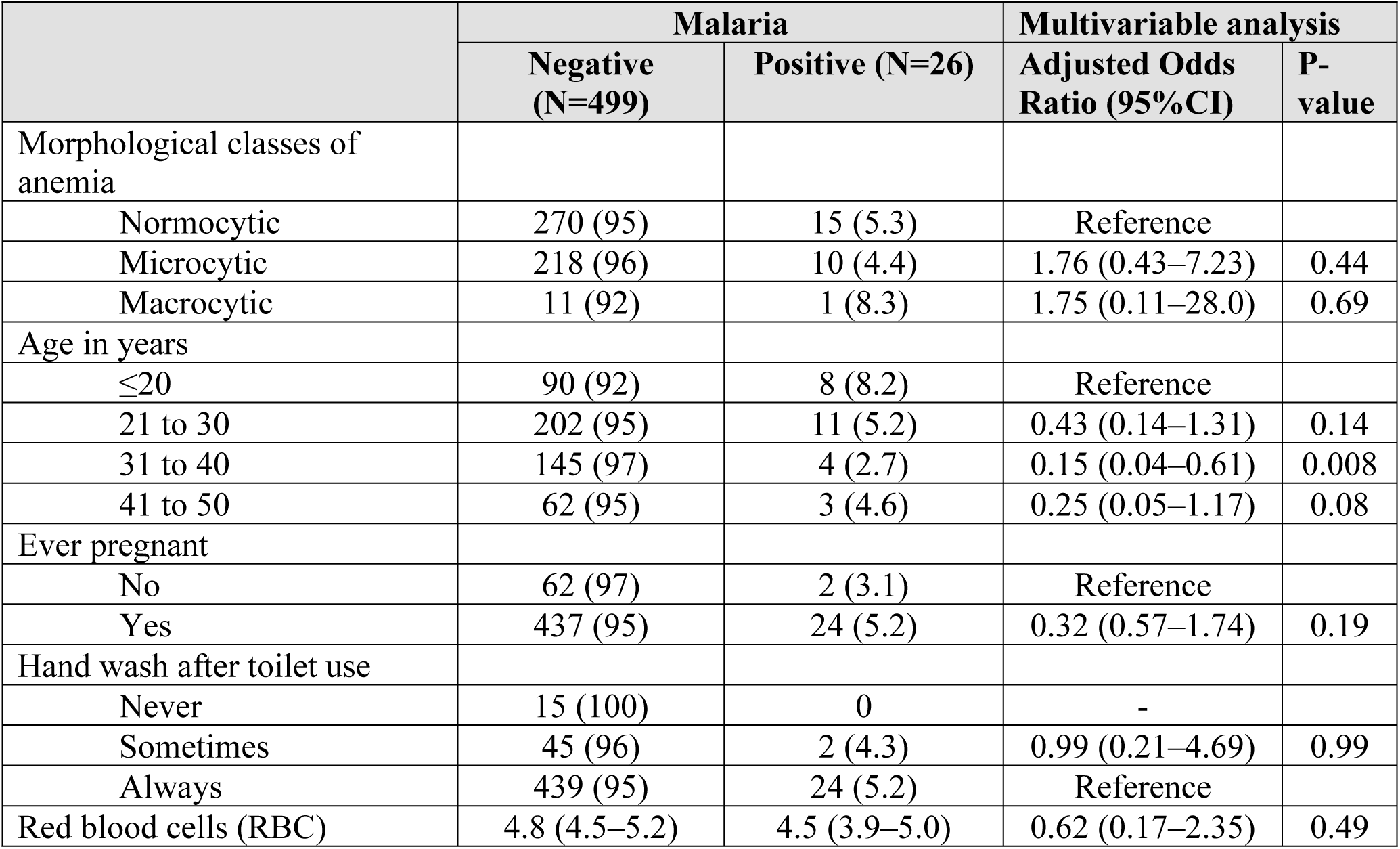
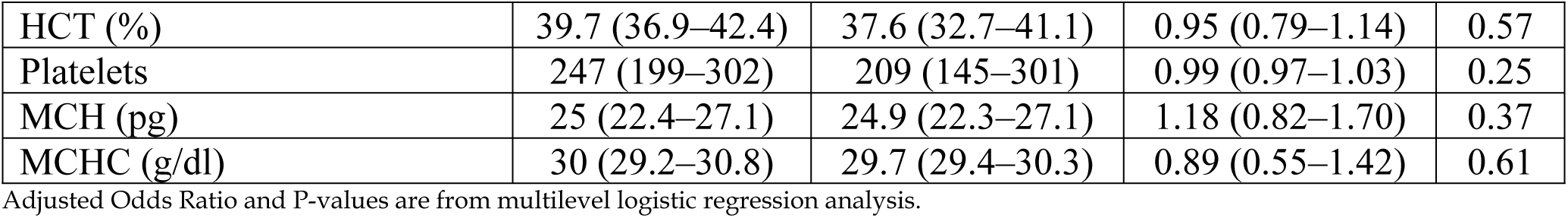
Analysis of the association between the morphological classes of anemia, with malaria, among women of reproductive age.

In the univariate analysis, Microcytic (COR 0.56 (95%CI 0.25‒1.27)) and macrocytic (COR 0.88 (95%CI 0.10‒7.33)) compared to normocytic were not associated with STH. After adjusting for potential confounders, Microcytic (aOR 0.51 (95%CI 0.13‒2.01)) and macrocytic (aOR 0.72 (95%CI 0.06‒9.39)) compared to normocytic were not associated with STH as shown in **Table 4**.

**Table 4.** Analysis of the association between the blood morphological change, with soil transmitted Helminthes among women of reproductive age.

|  | Soil Transmitted Helminthes (STH) |  | Multivariable analysis |  |
| --- | --- | --- | --- | --- |
|  | No (N=495) | Yes(N=30) | Adjusted Odds Ratio (95%CI) | P-value |
| Morphological classes of anemia |  |  |  |  |
| Normocytic | 265 (93) | 20 (7.0) | Reference |  |
| Microcytic | 219 (96) | 9 (4.0) | 0.51 (0.13–2.01) | 0.34 |
| Macrocytic | 11 (92) | 1 (8.3) | 0.72 (0.06–9.39) | 0.80 |
| Age in years |  |  |  |  |
| ≤20 | 94 (96) | 4 (4.1) | Reference |  |
| 21 to 30 | 199 (93) | 14 (6.6) | 1.32 (0.36–4.88) | 0.67 |
| 31 to 40 | 138 (93) | 11 (7.4) | 1.43 (0.36–5.68) | 0.62 |
| 41 to 50 | 64 (99) | 1 (1.5) | 0.24 (0.02–2.53) | 0.24 |
| Ever pregnant |  |  |  |  |
| No | 62 (97) | 2 (3.1) | Reference |  |
| Yes | 433 (94) | 28 (6.1) | 0.63 (0.11–3.44) | 0.59 |
| Hand wash after toilet use |  |  |  |  |
| Never | 13 (87) | 2 (13) | 2.60 (0.48–14.1) | 0.27 |
| Sometimes | 45 (96) | 2 (4.3) | 1.08 (0.24–4.95) | 0.92 |
| Always | 437 (94) | 26 (5.6) | Reference |  |
| Red blood cells (RBC) | 4.8 (4.4–5.2) | 4.7 (4.6–5.1) | 0.85 (0.22–3.32) | 0.82 |
| HCT (%) | 39.6 (36.9–42.3) | 40.6 (38.0–43.6) | 1.01 (0.84–1.21) | 0.95 |
| Platelets | 245 (196–299) | 265 (219–312) | 1.00 (0.98–1.02) | 0.11 |
| MCH (pg) | 24.9 (22.4–27.0) | 25.6 (22.7–27.3) | 0.98 (0.67–1.44) | 0.94 |
| MCHC (g/dl) | 30 (29.2–30.7) | 30.4 (29.4–30.8) | 0.99 (0.59–1.66) | 0.98 |

Adjusted Odds Ratio and P-values are from multilevel logistic regression analysis.

Normocytic anemia was not associated with Schistosomiasis in the univariate analysis (COR 1.27 (95%CI 0.51‒3.17)). After adjusting for age and pregnant status, Normocytic anemia was not associated with Schistosomiasis (aOR 1.37 (95%CI 0.54‒3.47)). Microcytic anemia was not associated with Schistosomiasis in the univariate analysis (COR 0.86 (95%CI 0.35‒2.15)) and after adjusting for age and pregnant status (aOR 0.78 (95%CI 0.31‒1.97)). No Macrocytic anemia case had Schistosomiasis **(Table 5).**

**Table 5.** Analysis of the association between the morphological classes of anemia, with schistosomiasis, Malaria and soil transmitted Helminthes (STH) among women of reproductive age.

|  | Schistosomiasis |  | Malaria |  | STH |  |
| --- | --- | --- | --- | --- | --- | --- |
|  | Odds Ratio (95%CI) | P-value | Odds Ratio (95%CI) | P-value | Odds Ratio (95%CI) | P-value |
| **Normocytic** |  |  |  |  |  |  |
| Bivariate analysis | 1.27 (0.51–3.17) | 0.60 | 1.15 (0.51–2.58) | 0.73 | 1.72 (0.78–3.77) | 0.18 |
| Adjusted for age & pregnancy | Adjusted Odds Ratio (95% CI) |  | Adjusted Odds Ratio (95% CI) |  | Adjusted Odds Ratio (95% CI) |  |
| Normocytic | 1.37 (0.54–3.47) | 0.50 | 1.14 (0.50–2.59) | 0.75 | 1.70 (0.77–3.76) | 0.19 |
| Age in years | 0.90 (0.84–0.97) | 0.007 | 0.93 (0.88–0.99) | 0.01 | 0.97 (0.92–1.02) | 0.22 |
| Ever pregnant | 2.04 (0.50–8.32) | 0.32 | 2.80 (0.57–13.8) | 0.21 | 2.37 (0.49–11.5) | 0.28 |
| **Microcytic** |  |  |  |  |  |  |
| Bivariate analysis | 0.86 (0.35–2.15) | 0.75 | 0.81 (0.36–1.84) | 0.62 | 0.57 (0.25–1.27) | 0.17 |
| Adjusted for age & pregnancy | Adjusted Odds Ratio (95% CI) |  | Adjusted Odds Ratio (95% CI) |  | Adjusted Odds Ratio (95% CI) |  |
| Microcytic | 0.78 (0.31–1.97) | 0.59 | 0.80 (0.35–1.84) | 0.60 | 0.56 (0.25–1.27) | 0.17 |
| Age in years | 0.90 (0.84–0.97) | 0.007 | 0.93 (0.88–0.99) | 0.01 | 0.97 (0.92–1.02) | 0.21 |
| Pregnant | 2.07 (0.51–8.42) | 0.31 | 2.76 (0.56–13.6) | 0.21 | 2.39 (0.49–11.6) | 0.28 |
| **Macrocytic** |  |  |  |  |  |  |
| Bivariate analysis | - |  | 1.66 (0.20–13.9) | 0.64 | 1.08 (0.13–8.92) | 0.94 |
| Adjusted for age & pregnancy | Adjusted Odds Ratio (95% CI) |  | Adjusted Odds Ratio (95% CI) |  | Adjusted Odds Ratio (95% CI) |  |
| Macrocytic | - | - | 2.23 (0.25–19.5) | 0.47 | 1.21 (0.14–10.1) | 0.86 |
| Age in years | 0.91 (0.84–0.97) | 0.008 | 0.93 (0.88–0.98) | 0.01 | 0.97 (0.92–1.02) | 0.24 |
| Pregnant | 2.11 (0.52–8.52) | 0.30 | 2.93 (0.60–14.4) | 0.19 | 2.53 (0.53–12.2) | 0.25 |

-; No data because No Macrocytic anemia cases had Schistosomiasis, Adjusted Odds Ratio and P-values are from multilevel logistic regression analysis.

## Discussion

This study aimed at assessing the prevalence of different morphological classes of anemia among women of reproductive age with schistosomiasis, soil-transmitted helminthes (STH), and malaria infections in Kwale County.

In regard to the morphological classes of anemia among the study participants, normocytic anemia was the most common with a prevalence of 54.3% followed by microcytic anemia at 43.4% and the least being macrocytic at 2.3% respectively. This study agrees with a study that was conducted in Uganda among lactating mothers which found that 41.5% of the study participants in Gulu had normocytic anemia, 30.8% had microcytic while only 3.1% had macrocytic anemia (26). Given the relatively young population of our study participants, this is expected since macrocytic anemia is generally found more commonly among the elderly (27). Anemic conditions may be attributed to heavy menstrual flow by many WRA, or heavy bleeding during delivery or may be injuries sustained during a woman’s daily chores (28). Other factors that may also contribute to anemia may include chronic diseases, renal malignant diseases, iron deficiency anemia, intestinal parasitosis, and malnutrition (7,25,27,29). These findings contrast with findings from another study that identified microcytic hypochromic anemia as the most common type among lactating women in Ethiopia (30). However, this study did not look at lactating status of the women under study. Furthermore, this study found that schistosomiasis was more prevalent among women with normocytic anemia while malaria and STH were more prevalent among women with macrocytic anemia. Interestingly, no women with macrocytic anemia were infected with schistosomiasis, indicating a potential protective or unrelated factor between this infection and macrocytic anemia. This is due to the fact that Plasmodium species and hookworms deplete the red blood cells of the host decreasing their numbers (30).

The distribution of morphological classes of anemia in different age groups was significantly different where normocytic and microcytic anemia cases were the most common at 21 to 30 and reduced with increase in age. This is consistent with other studies which have shown that normocytic and microcytic anemia is more prevalent among the younger population than in older populations (7). Our study participants ranged from 15 to 50 years of age which is considered a relatively young population. It is expected that, in the next few decades, as the population ages, more macrocytic anemias will be observed (7). There were more cases of the different morphological classes of anemia among WRA that had ever been pregnant than those that had never been pregnant. The difference here was significant and these results ascertain the fact that pregnant women are generally at a higher risk of anemia than non-pregnant women. Those that were not pregnant were more affected than those that were pregnant during the study. Several factors could account for this observation. For example, a study done among WRA across low and middle income countries found that non-pregnant women with a history of abortion were more likely to have anemia than those who did not (31). Further, those with multipara and grandpara were more likely to be anemic than those who had single births or none (31,32). Interestingly, there were significantly more cases of normocytic, microcytic, and macrocytic anemia among WRA that always washed hands than those that sometimes washed who were, in turn, significantly more than those who never washed hands after toilet use. The same trend was observed among WRA that always, sometimes, and never washed hands before preparing food or after helping a child defecate although the difference here was not significantly different. It is not clear what the connection could be between morphological classes of anemia and hand washing. A possible explanation would be that, from a hygiene perspective, those who always wash hands would seem to be more hygienic and therefore we would expect even infections with STH and malaria would be lower compared to the others. Logically, this would mean lower cases of anemia and therefore higher cases of normocytic women of reproductive age than either microcytic or macrocytic. Interestingly, the prevalence of parasitic infections among the WRA in this study, as reported previously (12) was low.

In terms of laboratory parameters, there were significantly more cases of normocytic and macrocytic anemia among WRA without anemia based on severity (i.e. no anemia based on hemoglobin concentration) and the cases decreased with increase in severity such that none anemia cases were more than mild anemia cases which in turn were more than moderate anemia cases and finally even less for severe anemia. These results agree with those of another study done in Uganda among lactating mothers which found a similar trend (26). However, this trend was not observed for microcytic anemia.

The median number of white blood cells was more among cases of macrocytic anemia followed by microcytic anemia and least for normocytic anemia. However, the difference here was not significant. The median number of red blood cells was higher for WRA with microcytic anemia than for normocytic and macrocytic. The difference was significant. However, these were within the normal range with the exception of the microcytic cases where there was a slight increase in the RBC numbers. Normal RBC reference numbers are 3.8 to 5.2 X 10^6^ cells per microliter (33). The median HCT was significantly higher among macrocytic than normocytic which was higher than microcytic anemia. HCT is a calculated value from the hematology analyzer and may not be as useful in assessing anemia as compared to hemoglobin (33). The median platelets numbers were significantly higher for macrocytic, followed by microcytic followed by normocytic. The primary function of platelets is to prevent and also stop bleeding. In a study of experimental mice, platelet production and count increased in iron deficiency anemia. Concurrently, their production and count correlated inversely with the level of hemoglobin (34) indicating the interconnectedness of platelet counts with anemia. The medians for MCH and MCHC were significantly higher for macrocytic followed by normocytic and least being microcytic. The MCH is the actual weight of hemoglobin while MCHC is the actual concentration of hemoglobin (33) and both depend on the cell size thereby making the observation seen here quite normal.

After adjusting for potential confounders, microcytic anemia compared to normocytic anemia were not associated with schistosomiasis. However, after adjusting for age, there were significant association of normocytic, microcytic, and macrocytic anemia with schistosomiasis but not significant after adjusting for pregnancy. In addition, after adjusting for potential confounders, microcytic and macrocytic anemia compared to normocytic anemia were not associated with malaria infection. However, after adjusting for age, normocytic, microcytic, and macrocytic anemia were significantly associated with malaria infection but not after adjusting for pregnancy. These findings suggest that age but not pregnancy is an important factor in the analysis of various morphological classes of anemia associated with schistosomiasis and malaria. However, this is not the case for STH infections. After adjusting for potential confounders, microcytic and macrocytic anemia compared to normocytic anemia were not associated with STH infections. Additionally, there was no association of normocytic, microcytic, and macrocytic anemia with STH infection even after adjusting for age and for pregnancy. Literature has shown that chronic diseases – including chronic parasitic infections – account for most anemia (3,35–37). Given that the prevalence and intensity of parasitic infections in this population were low as reported elsewhere (12) which were contributed by SBD, CBD, and use of improved WASH factors, it is possible that we were dealing with acute infections which might explain the lack of association of parasitic infections with the various morphological classes of anemia observed in this study. However, further studies are needed to determine if there may be other factors that might have contributed to the high prevalence of normocytic and microcytic classes of anemia in this population.

This study had several limitations. First, the Kato Katz method for determining STH infections which was used in this study is not sensitive enough, especially in low prevalence settings such as that found here. A new, more sensitive, technique known as FLOTAC (38) is recommended. Second, the single day urine may have led to missing some positive cases of *S. haematobium* considering the low prevalence of this infection in the study area. Third, considering the low prevalence of the infections among the WRA with different morphological classes of anemia, a much larger sample size would be ideal for a more reasonable comparison of the results, but this was limited by the resources available at the time of the study.

In conclusion the study showed that the prevalence of malaria, schistosomiasis, and STH infections among WRA with different morphological classes of anemia in the sub-counties under study were low. The study also revealed high prevalence of normocytic and microcytic anemia but low prevalence of macrocytic anemia among WRA in the area under study. Clearly, the infections observed did not contribute to the different morphological classes of anemia in Matuga and Kinango sub-Counties. Consequently, there is need for further studies to determine other factors possible associated with the anemia in this region that will inform the mechanisms and eventually the best way to tackle this high prevalence of the anemia observed.

## Data availability

The data of this work is available at https://doi.org/10.5281/zenodo.17132628.

## Acknowledgment

We wish to thank all the participants for accepting to be part of this research. We are grateful to the community health volunteers, nurses, and public health officers that were involved in one way or another in the accomplishment of this work.

## Author contribution

Funding acquisition; VTJ

Conceptualization; LN and VTJ

Sample and data collection; LN, VTJ, JHK, FM, LK; CO, CM, JM, JK, HK, and ZN

Supervision of the work; VTJ, JHK, and FM

Formal analysis; LN, MN, VTJ, JHK, and FM

Writing of initial draft; LN

Revising and editing; All authors.

Approval of final draft for submission; All authors.

## REFERENCES

1. DeMaeyer E, Adiels-Tegman M. The prevalence of anaemia in the world. World Heal Stat Q. 1985;38(3):302–16.

2. Warner MJ, Kamran MT. Iron Deficiency Anemia [Internet]. StatPearls Publishing. 2024. Available from: https://www.ncbi.nlm.nih.gov/books/NBK448065/

3. Gardner WM, Razo C, McHugh TA, Hagins H, Vilchis-Tella VM, Hennessy C, et al. Prevalence, years lived with disability, and trends in anaemia burden by severity and cause, 1990–2021: findings from the Global Burden of Disease Study 2021. Lancet Haematol. 2023;10(9):e713–34.

4. Farashi S, Harteveld CL. Molecular basis of α-thalassemia. Blood Cells, Mol Dis [Internet]. 2018;70(September 2017):43–53. Available from: 10.1016/j.bcmd.2017.09.004

5. Navya K., Prasad K, Singh BMK. Analysis of red blood cells from peripheral blood smear images for anemia detection: a methodological review. Med Biol Eng Comput [Internet]. 2022;60(9):2445–62. Available from: 10.1007/s11517-022-02614-z

6. Hamid M, Naz A, Alawattegama LH, Steed H. The Prevalence of Anaemia in a District General Hospital in the United Kingdom. Cureus. 2021;13(5).

7. Li X, Chen Q, Yang X, Li D, Du C, Zhang J, et al. Erythrocyte parameters, anemia conditions, and sex differences are associated with the incidence of contrast-associated acute kidney injury after coronary angiography. Front Cardiovasc Med. 2023;10(August):1–12.

8. Omer A, Hailu D, Nigusse G, Mulugeta A. Magnitude and morphological types of anemia differ by age among under five children: A facility-based study. Heliyon [Internet]. 2022;8(9):e10494. Available from: 10.1016/j.heliyon.2022.e10494

9. Colley DG, Bustinduy AL, Secor WE, King CH. Human schistosomiasis. Lancet. 2014;368(9541):2253–64.

10. Cando LFT, Perias GAS, Tantengco OAG, Dispo MD, Ceriales JA, Girasol MJG, et al. The Global Prevalence of Schistosoma mansoni, S. japonicum, and S. haematobium in Pregnant Women: A Systematic Review and Meta-Analysis. Trop Med Infect Dis. 2022 Nov;7(11):354.

11. Kihara JH, Kutima HL, Ouma J, Churcher TS, Changoma JM, Mwalisetso MA, et al. Urogenital schistosomiasis in women of reproductive age and pregnant mothers in Kwale County, Kenya. J Helminthol. 2015 Jan;89(1):105–11.

12. Jeza VT, Mutuku F, Kaduka L, Mwandawiro C, Masaku J, Okoyo C, et al. Schistosomiasis, soil transmitted helminthiasis, and malaria co-infections among women of reproductive age in rural communities of Kwale County, coastal Kenya. BMC Public Health. 2022 Jan;22(1):136.

13. Halliday KE, Oswald WE, McHaro C, Beaumont E, Gichuki PM, Kepha S, et al. Community-level epidemiology of soiltransmitted helminths in the context of school-based deworming: Baseline results of a cluster randomised trial on the coast of Kenya. PLoS Negl Trop Dis [Internet]. 2019;13(8):1–22. Available from: 10.1371/journal.pntd.0007427

14. Osazuwa F, Ayo OM, Imade P. A significant association between intestinal helminth infection and anaemia burden in children in rural communities of Edo state, Nigeria. N Am J Med Sci. 2011;3(1):30–4.

15. Grau-Pujol B, Gandasegui J, Escola V, Marti-Soler H, Cambra-Pellejà M, Demontis M, et al. Single-Nucleotide Polymorphisms in the Beta-Tubulin Gene and Its Relationship with Treatment Response to Albendazole in Human Soil-Transmitted Helminths in Southern Mozambique. Am J Trop Med Hyg. 2022;107(3):649–57.

16. Ng’etich AI, Amoah ID, Bux F, Kumari S. Anthelmintic resistance in soil-transmitted helminths: One-Health considerations. Parasitol Res [Internet]. 2024;123(1). Available from: 10.1007/s00436-023-08088-8

17. (DNMP) D of NMP, ICF. Kenya Malaria Indicator Survey 2020. 2020; (Nairobi Kenya and Rockville Maryland): DNMP and ICF.

18. Chaparro CM, Suchdev PS. Anemia epidemiology, pathophysiology, and etiology in low- and middle-income countries. Ann N Y Acad Sci. 2019 Aug;1450(1):15–31.

19. Mahmoud AAF, Woodruff AW. Mechanisms Ivolved in the Anaemia of Schistosomiasis. Trans R Sociaty Trop Med Hyg. 1972;66(1):75–84.

20. Butler SE, Muok EM, Montgomery SP, Odhiambo K, Mwinzi PMN, Secor WE, et al. Mechanism of anemia in Schistosoma mansoni-infected school children in Western Kenya. Am J Trop Med Hyg. 2012 Nov;87(5):862–7.

21. Kassebaum NJ, Jasrasaria R, Naghavi M, Wulf SK, Johns N, Lozano R, et al. A systematic analysis of global anemia burden from 1990 to 2010. Blood. 2014 Jan;123(5):615–24.

22. Salgado C, Ayodo G, Macklin MD, Gould MP, Nallandhighal S, Odhiambo EO, et al. The prevalence and density of asymptomatic Plasmodium falciparum infections among children and adults in three communities of western Kenya. Malar J. 2021 Sep;20(1):371.

23. Nyamu GW, Kihara JH, Oyugi EO, Omballa V, El-Busaidy H, Jeza VT. Prevalence and risk factors associated with asymptomatic Plasmodium falciparum infection and anemia among pregnant women at the first antenatal care visit: A hospital based cross-sectional study in Kwale County, Kenya. PLoS One. 2020;15(10):e0239578.

24. Katz N, Chaves A, Pellegrino J. A simple device for quantitative stool thick-smear technique in Schistosomiasis mansoni. Rev Inst Med Trop Sao Paulo. 1972;14(6):397–400.

25. Buttarello M. Laboratory diagnosis of anemia: are the old and new red cell parameters useful in classification and treatment, how? Int J Lab Hematol. 2016;38:123–32.

26. Clinton O, Micheal K, Angella KN, Mary M, Mugume M, Muwanguzi E, et al. Anaemia, Morphological Classification and Its Associated Risk Factors Among Lactating Mothers at Mbarara City Council Health Centre IV, Southwestern Uganda. J Blood Med. 2022;13(August):473–81.

27. Nagao T, Hirokawa M. Diagnosis and treatment of macrocytic anemias in adults. J Gen Fam Med. 2017;18(5):200–4.

28. Pita-Rodríguez GM, Basabe-Tuero B, Díaz-Sánchez ME, Alfonso-Sagué K, Álvarez AMG, Montero-Díaz M, et al. Prevalence of Anemia and Iron Deficiency in Women of Reproductive Age in Cuba and Associated Factors. Int J Environ Res Public Health. 2023;20(6):5110–20.

29. Yada TA, Dessie Y, Darghawth R, Wilfong T, Kure MA, Roba KT. Magnitude of Intestinal Parasitosis, Malnutrition, and Predictors of Anemia Among Nonpregnant Reproductive-Age Women Attending Healthcare Services in Olenchity General Hospital, Central Ethiopia. Front Trop Dis. 2021;2(July):1–11.

30. Feleke BE, Feleke TE. Pregnant mothers are more anemic than lactating mothers, a comparative cross-sectional study, Bahir Dar, Ethiopia. BMC Hematol. 2018;18:2.

31. Alem AZ, Efendi F, McKenna L, Felipe-Dimog EB, Chilot D, Tonapa SI, et al. Prevalence and factors associated with anemia in women of reproductive age across low- and middle-income countries based on national data. Sci Rep [Internet]. 2023;13(1):1–13. Available from: 10.1038/s41598-023-46739-z

32. Kumera G, Gedle D, Alebel A, Feyera F, Eshetie S. Undernutrition and its association with socio-demographic, anemia and intestinal parasitic infection among pregnant women attending antenatal care at the University of Gondar Hospital, Northwest Ethiopia. Matern Heal Neonatol Perinatol. 2018;4(1):1–10.

33. Doig K, Zhang B. A Methodical Approach to Interpreting the Red Blood Cell Parameters of the Complete Blood Count. Am Soc Clin Lab Sci. 2017;30(3):173–85.

34. Choi SI, Simone J V. Platelet production in experimental iron deficiency anemia. Blood [Internet]. 1973;42(2):219–28. Available from: 10.1182/blood.V42.2.219.219

35. Weigel MM, Calle A, Armijos RX, Vega IP, Bayas B V., Montenegro CE. The effect of chronic intestinal parasitic infection on maternal and perinatal outcome. Int J Gynecol Obstet. 1996;52(1):9–17.

36. Foy H, Nelson GS. Helminths in the Etiology of Anemia in the Tropics, with Special Reference to Hookworms and Schistosomes. Exp Parasitol. 1963;14:240–62.

37. Koukounari A, Estambale BBA, Kiambo Njagi J, Cundill B, Ajanga A, Crudder C, et al. Relationships between anaemia and parasitic infections in Kenyan schoolchildren: A Bayesian hierarchical modelling approach. Int J Parasitol. 2008;38(14):1663–71.

38. Utzinger J, Rinaldi L, Lohourignon LK, Rohner F, Zimmermann MB, Tschannen AB, et al. FLOTAC: a new sensitive technique for the diagnosis of hookworm infections in humans. Trans R Soc Trop Med Hyg. 2008;102(1):84–90.

